# Predicting the age of Alzheimer’s symptom onset from a blood test: evaluating a prognostic claim

**DOI:** 10.64898/2026.03.26.26349230

**Authors:** Philip S. Insel, Michael C. Donohue

**Author notes:** Corresponding author: Philip S. Insel, Department of Psychiatry and Behavioral Sciences, University of California, San Francisco, CA, USA.

## Abstract

**Objectives:** Plasma p-tau217 clocks have been reported to predict when Alzheimer’s symptoms will begin, from a single blood test. This would transform Alzheimer’s research and care. We evaluated the prognostic value of these models.

**Design:** Analysis of a published prognostic study using data from two prospective longitudinal cohorts.

**Setting:** Two observational research cohorts: the Knight Alzheimer Disease Research Center (Knight ADRC), a single US centre, and the Alzheimer’s Disease Neuroimaging Initiative (ADNI), a multicentre study in the US and Canada.

**Participants:** 345 ADNI participants with longitudinal plasma %p-tau217 used to develop the clocks; 200 ADNI participants who were cognitively unimpaired (Clinical Dementia Rating 0) at blood draw; and 83 participants from both cohorts who progressed to symptomatic Alzheimer’s disease in the original report.

**Main Outcome Measures:** Age at symptom onset; time from %p-tau217 positivity to onset; time to onset from blood draw.

**Results:** The clock predictor, estimated age at %p-tau217 positivity, is age at blood draw minus estimated time since positivity, so age is shared by the predictor and the outcome, age at symptom onset. Separated from age, the clock’s %p-tau217 component explained essentially no variance (adjusted R² −0.02 to 0.02). Age alone explained 0.54 to 0.71, compared with 0.34 to 0.61 reported for the clock. Raw %p-tau217 with age outperformed the clock in every analysis. For time from positivity to onset, the reported shorter interval at older positivity ages was reproduced when the %p-tau217 component was replaced with a random number. In survival analysis, estimated age at positivity was associated with symptom onset age (hazard ratio 0.87, 95% confidence interval 0.81 to 0.95), but not when onset was measured from blood draw, removing the shared age component (1.02, 0.96 to 1.09).

**Conclusions:** The clock claims are unsupported. The reported accuracy comes from age shared between predictor and outcome, not from %p-tau217. Because timing is estimated only in known progressors, the model does not attempt to identify *if* an individual will progress, so it cannot answer the key question of *when* they will progress. Predictions from these models are unsupported for clinical or consumer use.

## Introduction

The Alzheimer disease field has advanced markedly in recent years, in large part due to blood-based biomarkers. Plasma p-tau217, in particular, identifies underlying amyloid and tau pathology with high accuracy,^1,2^ at a fraction of the cost of PET or cerebrospinal fluid testing, and is now moving into clinical use. Pathology that once required a PET scan or a lumbar puncture to detect can now be identified from a blood sample.

With detection increasingly feasible, the question is now how prognostic a blood test can be: whether it can forecast an individual’s disease trajectory, and above all when symptoms will appear. Disease clock models^3,4^ have become a leading approach. From a single p-tau217 level, the model reads off how far along an assumed common trajectory a person has progressed, and converts that position to a time scale, estimating the age at which they became positive and the time remaining before symptoms. Their predictive power and simplicity come from one assumption: that every individual, once accumulation begins, progresses along the same trajectory at a consistent rate, so that a single measurement can be read as a point in time. First applied to amyloid PET, the approach has since been extended to plasma biomarkers, and such models now make up a rapidly growing literature.^3–8^

Applying disease clock models (SILA^3^ and TIRA^4^) to plasma %p-tau217, Petersen and colleagues^9^ estimate each individual’s age at %p-tau217 positivity, defined as age at blood draw minus the estimated time from positivity, and regress observed age at symptom onset on this estimate in individuals who progressed to symptomatic AD. On the basis of adjusted R² values of up to 0.6, they conclude that age at symptom onset can be estimated from a single blood test with a median error of less than four years. The report has drawn substantial international press coverage,^10,11^ the models are implemented in a publicly accessible web application that returns individual predictions of symptom onset,^12^ and, with plasma p-tau217 available direct to consumer, these predictions have a route to individual patients with no clinical oversight.

Predictive accuracy of this magnitude would genuinely transform Alzheimer research. The age at symptom onset could be estimated from a single blood measurement years in advance, onset anticipated for an individual patient, interventions timed accordingly, and trials enriched by predicted onset with substantial gains in efficiency.

Precisely because these implications are so large, the size of the reported effect warrants close scrutiny. An R² of 0.6 for a clinical outcome, from a single blood measurement, is large. Effects of this size have warranted a closer look before. In social neuroscience, implausibly strong brain–behavior correlations were traced to analyses that used the same data to select regions of interest and to estimate the correlation within them, a problem identified only after the approach had become standard practice.^13^

Associations of this kind often arise from design features that recur across fields, each of which inflates apparent associations. A shared component arises when the same variable contributes to both predictor and outcome, so that a portion of the explained variance is guaranteed independently of any real association. A bound arises when the outcome cannot take any value but is limited to a region fixed by the predictor, so that the constraint itself manufactures an association. Disease clocks may be prone to both, and we examine whether the reported associations arise from these features rather than from information in the biomarker.

In the analyses that follow, we examine each mechanism in turn. We test whether the reported accuracy persists when the biomarker is replaced by noise, whether it holds when the shared component is removed, and whether a simpler model using the raw plasma measurement directly performs comparably. Finally, we assess whether the data support the assumption of consistent accumulation on which the framework rests.

## Methods

### Participants

Participants in the progressors analyses were cognitively unimpaired at their initial clinical visit (CDR 0) and had at least one plasma %p-tau217 measure. The progressor analyses used data shared by the original authors: Knight Alzheimer Disease Research Center (Knight ADRC) and Alzheimer’s Disease Neuroimaging Initiative (ADNI) participants who progressed to symptomatic AD (CDR > 0). The survival analyses used contemporary ADNI data (downloaded 10 July 2026), including nonprogressors, and the common-trajectory analysis used the ADNI clock-development sample (n=345). As in the original report, clock estimation required at least two plasma %p-tau217 values within the range used to develop the clocks, separated by at least one year, and clock assignment required at least one in-range value.^9^

### Plasma %p-tau217

As in the original report, plasma %p-tau217, the ratio of phosphorylated to non-phosphorylated tau at position 217, was measured by liquid chromatography–mass spectrometry (C2N Diagnostics). The positivity threshold was 4.06, corresponding to an amyloid PET level of 20 centiloids, and the clocks were developed over the range 1.06 to 10.45.^9^

### Clock Estimation

TIRA estimates the time required to pass from one biomarker level to another by integrating the reciprocal of the modeled rate of accumulation.^4^ For plasma %p-tau217, individual rates were estimated with a linear mixed-effects model and related to the level at the midpoint of follow-up with a generalized additive model.^9^ SILA samples biomarker versus age data to establish the relationship between rate and level, and integrates it numerically by Euler’s method into a curve of biomarker level against time.^3^

For the progressor analyses, TIRA and SILA estimates were supplied by the authors of the original report; participants with more than one plasma sample were assigned the average of their estimates.

### Clinical Measures

Cognitive status was assessed with the Clinical Dementia Rating (CDR). Symptomatic AD was defined as a CDR global score greater than 0 with a diagnosis of an AD syndrome.^9^ For the survival analyses, participants were required to be cognitively unimpaired (CDR 0) at the qualifying blood draw. Onset was dated to the start of the interval in which impairment occurred, and it had to persist at the last assessment and occur after the estimated age at positivity. Participants who progressed before their estimated age at positivity, or to a non-AD syndrome, were censored at their last unimpaired assessment. Clinical syndrome was assessed using the CDR by experienced clinicians blinded to biomarker results.^9^

### Statistical Analysis

In each analysis, the aim was to separate the association attributable to biomarker signal from the association produced by two design features: a shared component, in which the same quantity enters both the predictor and the outcome, and a bound, in which the outcome is confined to a region fixed by the predictor. Full details of each analysis are given in the Supplementary Methods.

We decomposed the clock into age at blood draw, defined as age at the first plasma collection, and estimated time from %p-tau217 positivity (time from positivity). In the first set of analyses, age at symptom onset was regressed by ordinary least squares on age at blood draw, on time from positivity, and on the clock-estimated age at positivity, each separately, and the clock was compared with a model using age at blood draw and raw %p-tau217 directly, by adjusted R² and the Akaike information criterion.^14^ As a null comparison, time from positivity was replaced with a random value drawn uniformly over the same range, 10,000 times. These comparisons were repeated after excluding participants whose symptom onset preceded the blood draw.

In the second set, age was removed from the outcome by regressing time from blood draw to symptom onset on time from positivity, estimated by TIRA, by SILA, and as a random value; the analysis was repeated in participants already positive at the draw, for whom the bound places little constraint on time to onset. In the third set, time from positivity to symptom onset was regressed on estimated age at positivity, estimated in the same three ways; because the outcome contains the predictor, the two are negatively correlated by construction.^15^

In the fourth set, nonprogressors were included and interval-censored proportional hazards models (icenReg,^16^ as in the original report) were fit on two timescales: age, as in the original report, and time from the blood draw or from estimated positivity for participants not yet positive at the draw, which excludes immortal time before an event could be counted.^17,18^ Confidence intervals were obtained by bootstrap resampling, and individual prediction intervals from the fitted survival curves. Survival curves are shown until fewer than five participants remained under observation.

In the fifth set, we tested the assumption that individuals at the same %p-tau217 level progress at the same rate. In the ADNI clock-development sample (n=345), participant-specific rates of change were estimated from a linear mixed model and regressed on %p-tau217 level, using either the observed value at the first plasma collection or the model-based value at the midpoint of follow-up, with uncertainty propagated by resampling.

## Results

### Participants

Data provided by the authors comprised participants from the original report who were cognitively unimpaired at their first clinical assessment, developed cognitive impairment with an AD syndrome after estimated %p-tau217 positivity, and remained impaired with an AD syndrome at their last assessment: 59 (TIRA) and 61 (SILA) individuals from the Knight ADRC and 20 (TIRA) and 22 (SILA) from ADNI, described in Supplementary Table 2 of Petersen et al.^9^ Clinical follow-up began before the first plasma collection, and 11% to 15% of these participants had symptom onset before their blood draw (Knight ADRC: 7 of 61 for SILA and 8 of 59 for TIRA; ADNI: 3 of 22 for SILA and 3 of 20 for TIRA). Median clinical follow-up was 13.0 to 13.2 years in the Knight ADRC progressors (11 assessments) and 10.1 years in the ADNI progressors (10.5 assessments).

The ADNI survival sample comprised 200 participants who were cognitively unimpaired at the qualifying blood draw and had a SILA-estimated age at %p-tau217 positivity. Median age at blood draw was 74.1 years, 54.0% were female, median education was 16 years, 27.5% carried at least one *APOE* ε4 allele, and median follow-up from blood draw was 8.22 years. Thirty progressed to symptomatic AD after their estimated age at %p-tau217 positivity. Seven progressed before their estimated age at positivity and 30 to a non-AD diagnosis; both groups were censored at their last unimpaired assessment. The remaining 133 were unimpaired at their last assessment and were censored at that time. For the analysis with follow-up starting at the blood draw or estimated positivity, 64 participants whose follow-up ended before their estimated age at positivity were excluded, including the 7 who progressed before positivity, leaving 136 participants and 30 events. In 76 of these, follow-up began at the estimated age at positivity.

The analyses that follow model three outcomes: age at symptom onset, time from blood draw to onset, and time from %p-tau217 positivity to onset. Each analysis in the original report contains a shared component: the same quantity enters both the predictor and the outcome, so the two are correlated by construction. All three analyses are also restricted to individuals whose age at symptom onset was greater than their estimated age at positivity. This inclusion rule bounds the outcome, so it cannot take just any value and is confined to a region fixed by the predictor. The rule is the same for every outcome; only the quantity that sets the bound changes. Either feature alone is sufficient to manufacture an association, whether or not the biomarker carries any information. The shared component and the bound for each outcome are described in the sections that follow.

### Age at symptom onset: a shared component and a bound

In the original report, age at symptom onset was regressed on the clock’s estimated age at %p-tau217 positivity, defined as age at blood draw minus estimated time from positivity. Age at blood draw is therefore the shared quantity: it enters the predictor directly, and it is contained in the outcome, since age at onset equals age at blood draw plus time from blood draw to onset.

Age at blood draw also bounds the outcome. Onset could be observed only within a limited window of clinical follow-up, which began before the blood draw, so age at onset is confined to a fixed band around age at blood draw, from 5.7 years before to 16.4 years after blood draw in Knight ADRC (Figure 1).

**Figure 1.**
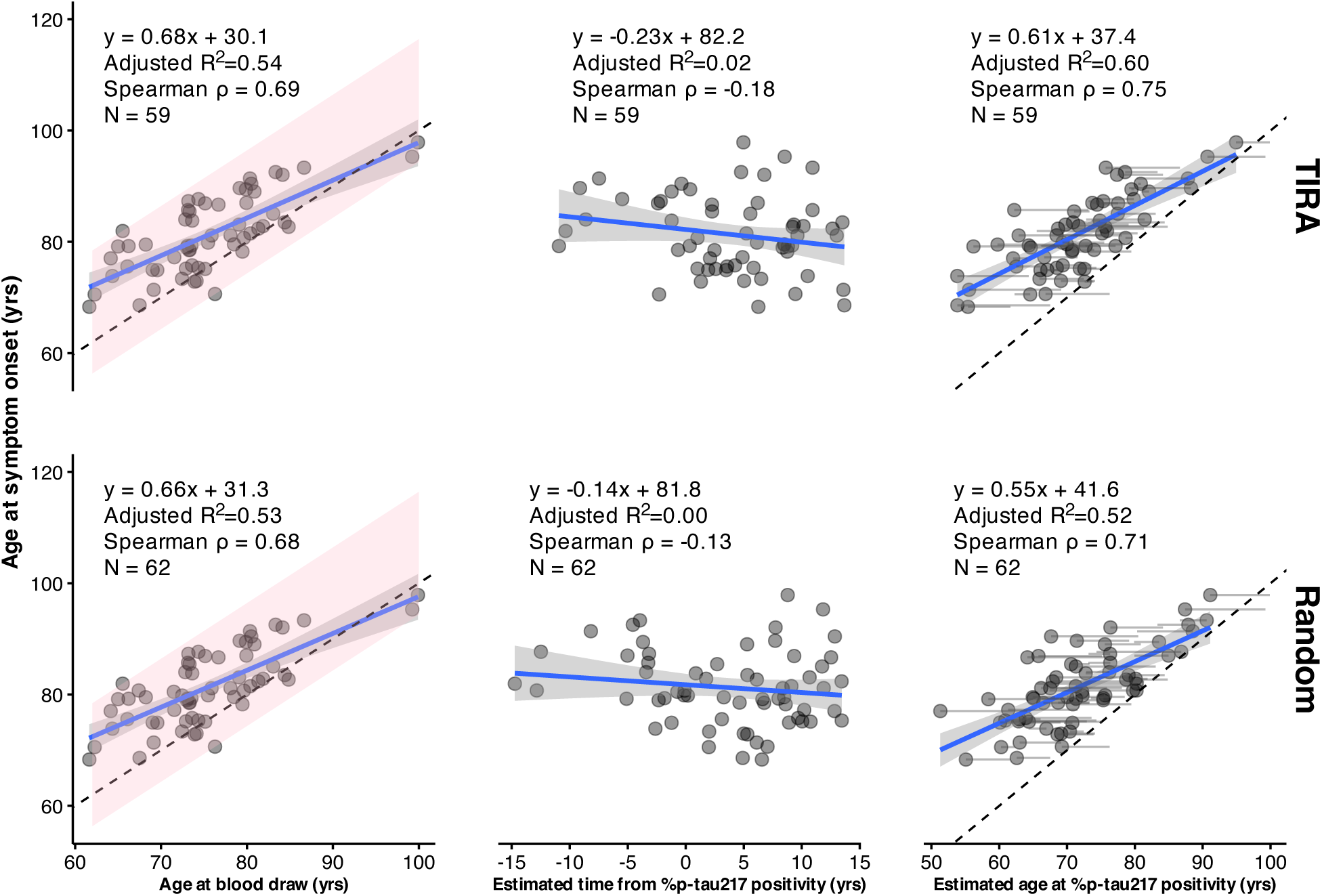
Relationships in Knight ADRC between the predicted outcome (age at symptom onset) and three predictors: age at blood draw (first plasma collection), estimated time from %p-tau217 positivity, and their difference, age at blood draw minus estimated time from %p-tau217 positivity (ie, estimated age at %p-tau217 positivity). Time from %p-tau217 positivity is estimated by the TIRA method (top row) or by randomly sampling from a uniform distribution over the observed range of estimates (bottom row); results for the SILA method are shown in Supplementary Figure S2. Panels include fitted linear regression estimates, summary statistics for each predictor, and a dashed identity line. To illustrate the contribution of time from %p-tau217 positivity (middle column), the rightmost panels show faint grey lines connecting each person’s age at blood draw to their estimated age at %p-tau217 positivity. The pink regions span the observed window of onset, from 5.7 years before to 16.4 years after the blood draw. Because onset was observed only within this window, the data are constrained to a fixed band around the identity line, and age at blood draw determines age at onset up to that interval.

The original report attributed adjusted R² values of 0.337 to 0.612 to the clock-estimated age at %p-tau217 positivity as a predictor of age at symptom onset.^9^ When the clock was decomposed into age at blood draw and estimated time from %p-tau217 positivity, the biomarker timing term alone explained essentially no variance in age at symptom onset in either cohort: adjusted R² = 0.02 (TIRA) and 0.01 (SILA) in the Knight ADRC cohort (Figure 1) and adjusted R² = −0.02 (TIRA) and 0.00 (SILA) in ADNI (Supplementary Figure S1). TIRA and SILA estimates in both cohorts are shown in Supplementary Figure S2. Age at blood draw alone explained far more: adjusted R² = 0.54 for both method samples in Knight ADRC, and adjusted R² = 0.71 (TIRA sample) and 0.61 (SILA sample) in ADNI.

Incorporating the biomarker timing term through the clock produced some improvement over age alone in Knight ADRC (adjusted R² = 0.60–0.61 vs 0.54; ΔR² = 0.06–0.07). However, this improvement was largely driven by participants whose blood was drawn after symptom onset (7 of 61 for SILA; 8 of 59 for TIRA). These draws came from people who were already cognitively impaired, whereas the analysis is premised on predicting time to symptoms in people who are cognitively unimpaired (CDR 0); for these participants, the clock recovers an onset that has already occurred rather than predicting one. When they were excluded, the clock’s advantage over age fell to less than 0.02 for both methods (SILA: adjusted R² = 0.55 for the clock vs 0.54 for age; TIRA: 0.56 for the clock vs 0.55 for age). In ADNI the clock performed worse than age alone (TIRA: adjusted R² = 0.34 for the clock vs 0.71 for age; SILA: 0.47 for the clock vs 0.61 for age) and worse than a random comparator (adjusted R² = 0.56, Supplementary Figure S1), and excluding participants with onset before the draw (3 in each sample) widened the gap (TIRA: 0.33 for the clock vs 0.83 for age; SILA: 0.47 for the clock vs 0.74 for age).

Under the design-constrained null, in which biomarker timing information was replaced by random values, the nominal Type I error rate of the clock was severely inflated. In Knight ADRC, 100% of simulated analyses produced statistically significant associations (p<0.05), despite the absence of biomarker information; in ADNI, 98.5% were significant. The median null adjusted R² ranged from 0.43 to 0.49, accounting for most of the observed predictive performance: 77 to 82% of the observed adjusted R² in the Knight ADRC analyses and 91 to 141% (greater than 100% indicates worse performance than the median null value) of the observed adjusted R² in ADNI. Thus, the magnitude of the reported associations is largely what would be expected from the study design alone, even when biomarker information is removed, and in the case of ADNI, worse than expected.

### Time from blood draw to symptom onset

Removing age at blood draw from the outcome, by modeling time from blood draw to symptom onset rather than age at onset, removes the shared quantity but not the bound. The bound now comes from estimated time from %p-tau217 positivity, because the inclusion rule requires the time of onset to exceed the time of positivity. A participant with a time from positivity of −5 years, predicted to become positive 5 years after the blood draw, could be included only if onset occurred more than 5 years after the draw. For every participant not yet positive at the draw, time from positivity therefore sets a floor on time to onset. We show this by regressing time from blood draw to symptom onset on time from positivity, a regression in which age at blood draw appears on neither side. Participants not yet positive at the draw are pushed into the upper left of Figure 2, and a negative slope is guaranteed whether or not the biomarker carries any information.

**Figure 2.**
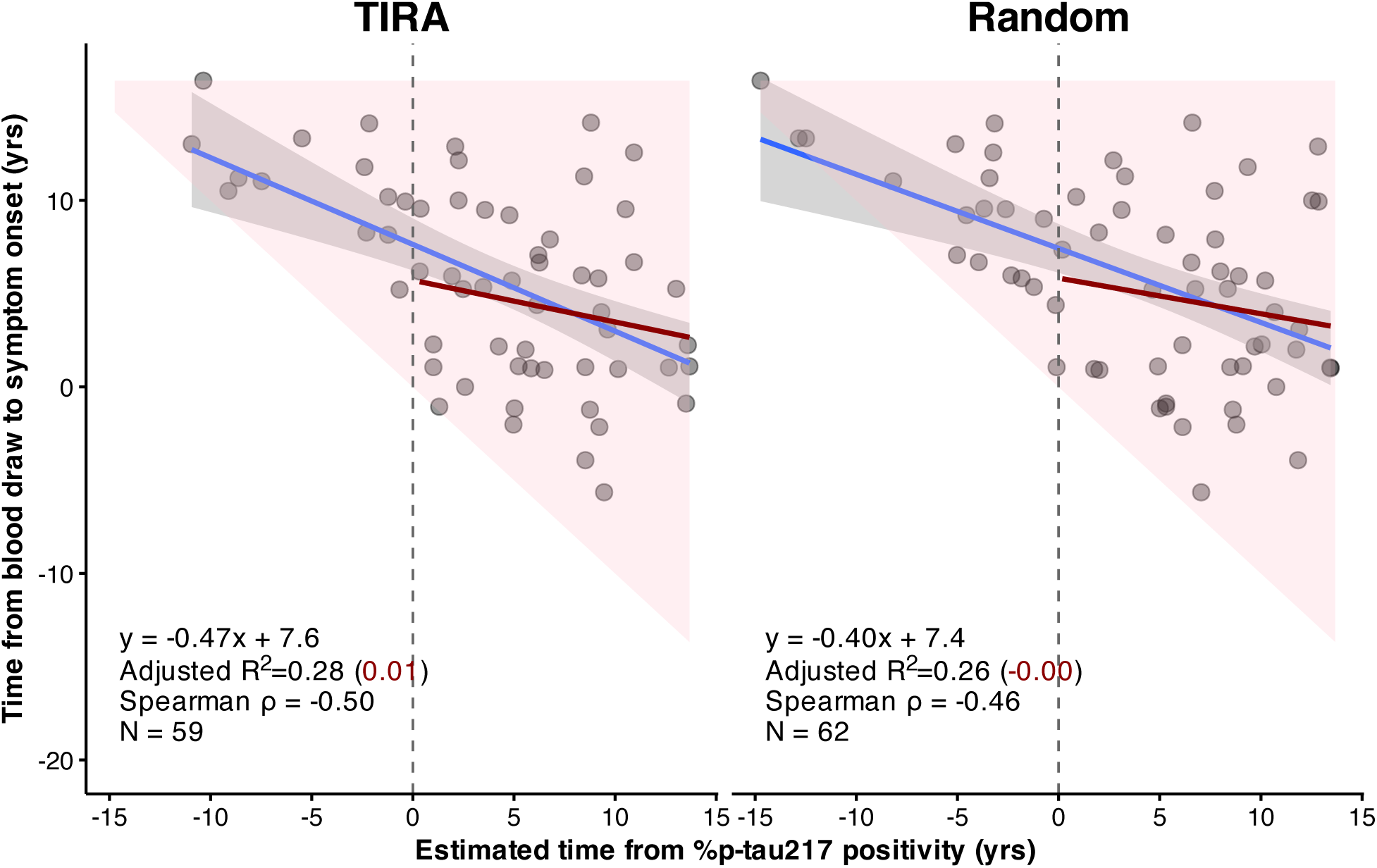
Time from blood draw to symptom onset versus estimated time from %p-tau217 positivity in Knight ADRC, with time from positivity estimated by the TIRA method (left) or drawn uniformly over the observed range of estimates (right); results for the SILA method and for ADNI are shown in Supplementary Figure S3. Negative values of estimated time from positivity indicate participants not yet positive at the blood draw (left of the dashed vertical line). The pink region shows the values permitted by the design: because onset must follow estimated positivity, time to onset cannot fall below the diagonal. Blue lines show linear fits in all participants, with 95% confidence bands; dark red lines show fits restricted to participants already positive at the blood draw. Each panel reports the regression equation, adjusted R² for all participants (restricted sample in parentheses), Spearman ρ, and N.

In Knight ADRC, time from positivity explained a substantial share of the variance in time to onset in the full sample, where the bound constrains participants not yet positive at the draw (adjusted R² 0.28 to 0.31), and a random comparator performed similarly (0.26). When the analysis was restricted to participants already positive at the blood draw, for whom the bound falls before the draw and has little effect, adjusted R² fell to 0.01 to 0.03 (Figure 2). In ADNI, adjusted R² was negative in the restricted sample for both methods (Supplementary Figure S3).

### Direct Use of %p-tau217

When %p-tau217 was entered into the model directly, as a raw level alongside age at blood draw, it outperformed the clock in every cohort and for every clock method (Figure 3). In ADNI, however, raw %p-tau217 added little beyond age at blood draw alone. Excluding participants whose blood was drawn after symptom onset widened the advantage of age and raw %p-tau217 over the clock in Knight ADRC: adjusted R² was 0.67 for age and raw %p-tau217 vs 0.55 for the clock (SILA) and 0.67 for age and raw %p-tau217 vs 0.56 for the clock (TIRA).

**Figure 3.**
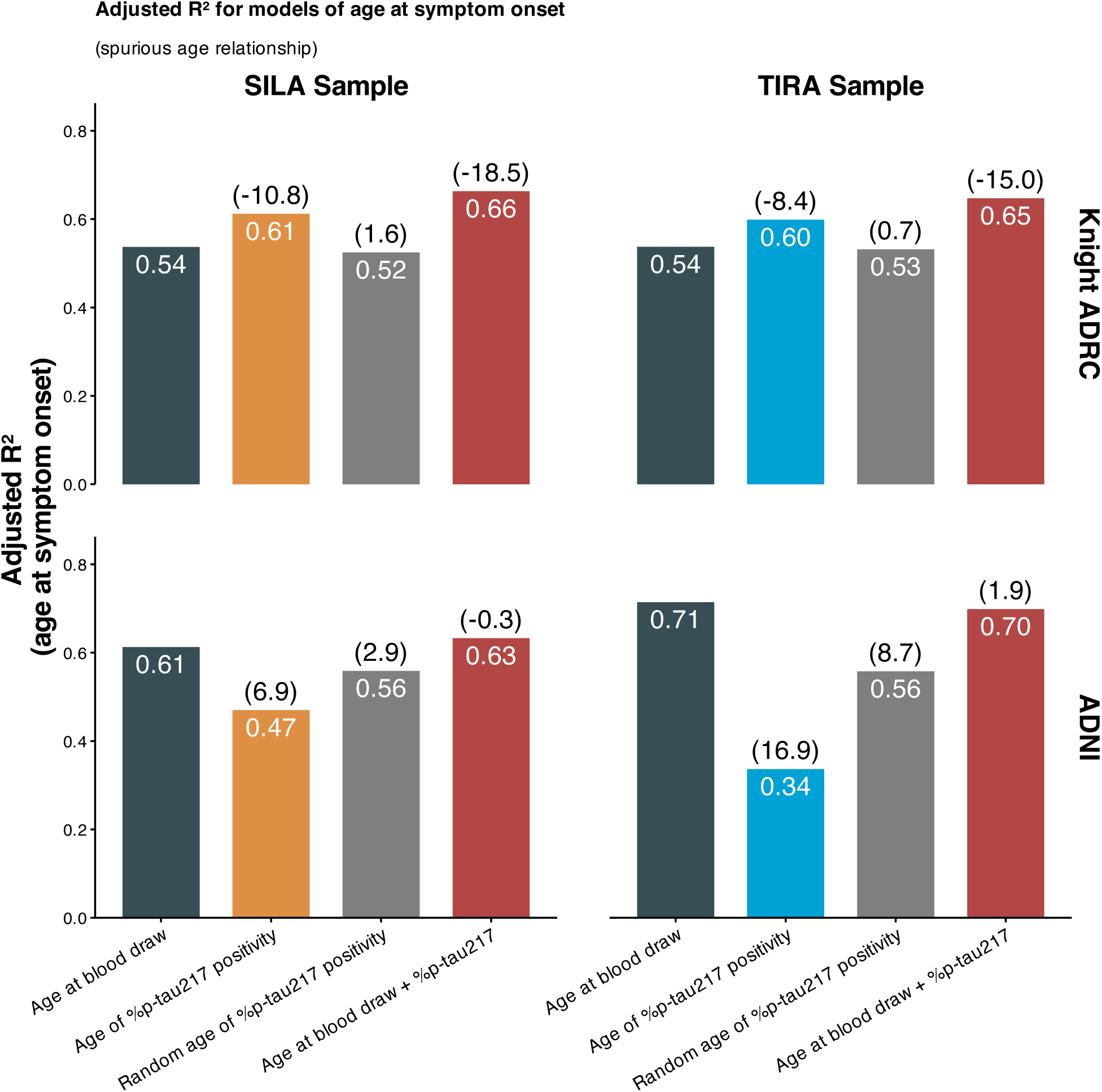
Model comparison across cohorts and clock methods. Adjusted R² for four models of age at symptom onset in Knight ADRC (top row) and ADNI (bottom row): age at blood draw alone (dark), the clock-derived age at %p-tau217 positivity (orange in the SILA column, blue in the TIRA column), a random comparator for age at %p-tau217 positivity from a single draw uniformly over the observed range (grey), and age at blood draw together with raw %p-tau217 level (red). Columns correspond to the samples fit with SILA and TIRA. Values inside bars give adjusted R²; values in parentheses above bars give the change in Akaike information criterion (ΔAIC) relative to the age-at-blood-draw model in the same panel, with negative values indicating better fit and positive values worse fit. Across cohorts, the clock-derived predictor does not improve on direct inclusion of age and raw %p-tau217.

### Relocation of the Bound: Analyses of the time from %p-tau217 positivity to symptom onset

The original report also regressed time from %p-tau217 positivity to symptom onset on estimated age at positivity. Here the shared quantity is the predictor itself: the outcome is age at onset minus estimated age at positivity, so the predictor enters the outcome with a negative sign, and the two are negatively correlated by construction. Estimated age at positivity also bounds the outcome. Because observed symptom onset in these cohorts falls between roughly 68 and 98 years, the duration can be at most 8 years for someone positive at 90 and must be at least 8 years for someone positive at 60 (shaded regions, Figure 4; both TIRA, SILA and both cohorts shown in Supplementary Figure S4). The reported finding that older age at positivity is associated with a shorter time to symptom onset therefore follows from the design, independent of any biomarker signal. Replacing the clock-derived estimate with age at blood draw minus a randomly generated time reproduced a negative association of similar magnitude to the SILA and TIRA estimates.

**Figure 4.**
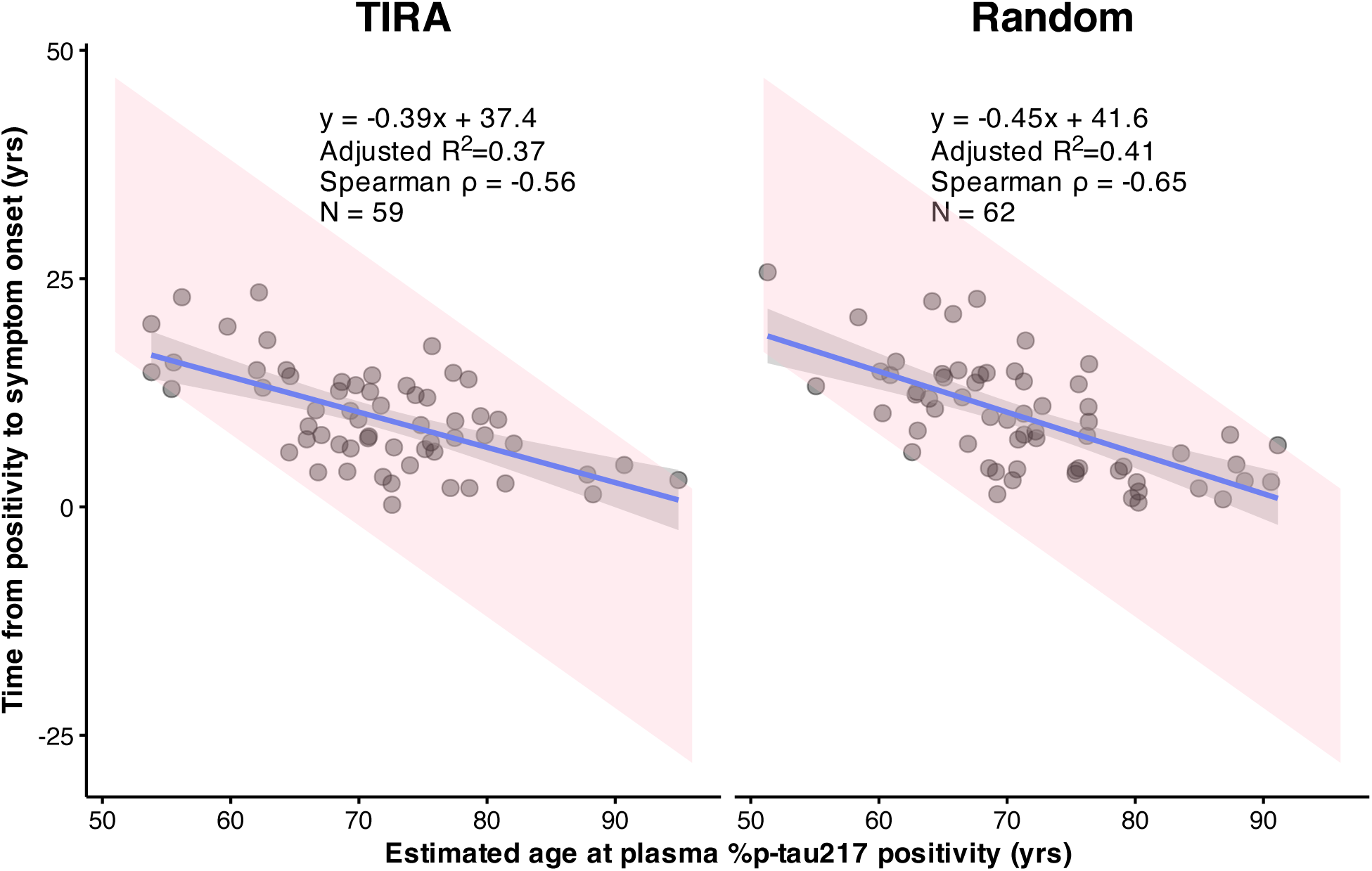
Time from estimated %p-tau217 positivity to symptom onset (age at symptom onset minus estimated age at positivity) versus estimated age at positivity in Knight ADRC, with age at positivity estimated by the TIRA method (left) or as age at blood draw minus a random time drawn uniformly over the observed range of estimates (right); results for the SILA method and for ADNI are shown in Supplementary Figure S4. Panels include fitted linear regression lines with 95% confidence bands, the regression equation, adjusted R², Spearman ρ, and N. The pink regions show the values possible given the observed range of ages at symptom onset (68 to 98 years): the older the estimated age at positivity, the shorter the duration can be.

### Survival Models on 2 Timescales: Age at symptom onset vs Time to symptom onset

Including nonprogressors, as in a survival analysis, might be expected to resolve these problems, since the sample is no longer restricted to those who progressed. On the age timescale used in the original report, however, the shared quantity remains: age at blood draw enters both the predictor, estimated age at positivity, and the outcome, age at symptom onset. The bound also remains, because onset is counted as an event only when it occurs after estimated age at positivity. In the ADNI survival sample (n=200; 30 events), a survival analysis with the shared quantity found estimated age at positivity was associated with symptom onset on the age timescale (HR 0.87, 95% CI 0.81 to 0.95, P<0.001, Figure 5a).

**Figure 5.**
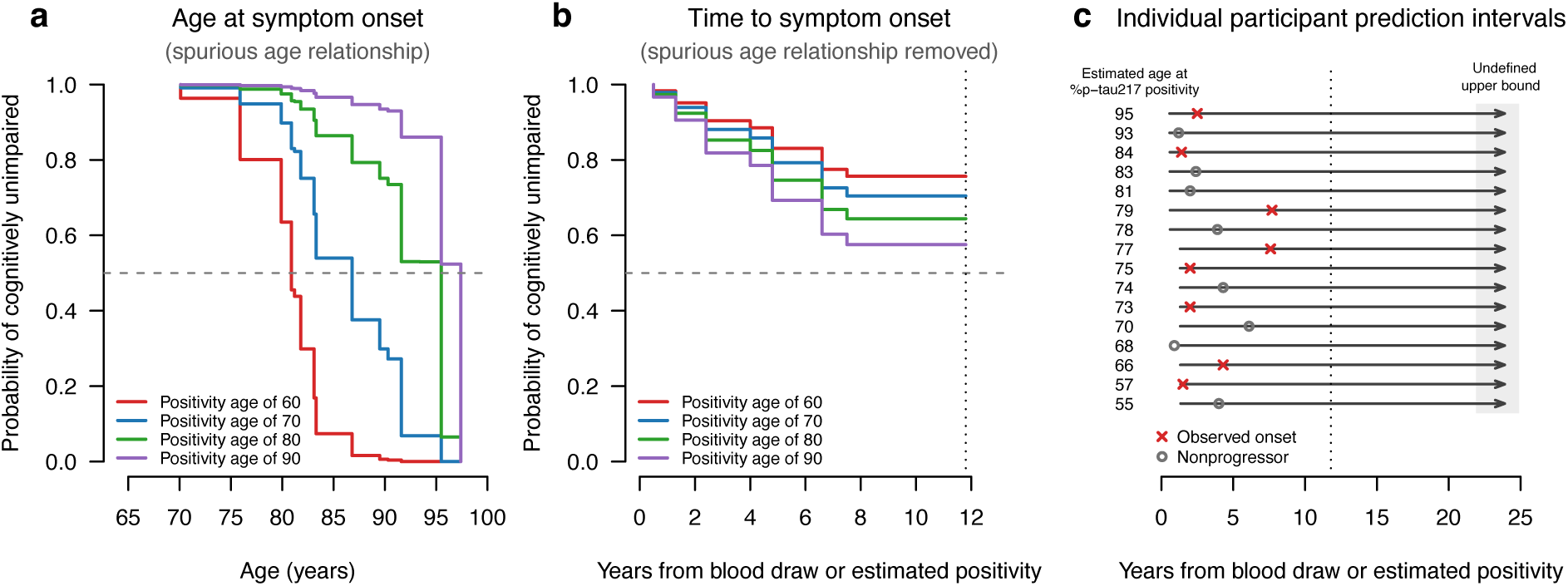
Survival models of symptom onset in the ADNI survival sample. Interval-censored proportional hazards models of estimated age at %p-tau217 positivity (SILA), fit (a) on the age timescale used in the original report (n=200; 30 events) and (b) on time from the blood draw, or from the estimated time of positivity for participants not yet positive at the draw (n=136; 30 events). Curves show predicted survival for estimated positivity ages of 60, 70, 80 and 90 years; the dashed horizontal line marks a survival probability of 0.5. In (b), curves are shown until fewer than five participants remained under observation (dotted vertical line, 11.8 years). (c) 95% prediction intervals for time to symptom onset from the model in (b) for 16 participants, eight with observed onset and eight without, chosen to span the range of estimated positivity ages. Each interval begins at its lower 2.5% limit; arrows extending to the shaded band indicate that no upper limit is reached within follow-up (dotted line). Crosses mark observed onset, and circles mark the last assessment of participants who remained unimpaired.

We then measured time to symptom onset from the blood draw, or from the estimated time of positivity for participants not yet positive at the draw. This removes the shared quantity, because age no longer appears in the outcome; it removes the bound, because no follow-up precedes estimated positivity; and it removes immortal time bias,^17,18^ because follow-up before the blood draw and before estimated positivity, when no event could be counted, is no longer included. On this timescale, estimated age at positivity was not associated with symptom onset (n=136; 30 events; HR 1.02, 95% CI 0.96 to 1.09, P=0.51; Figure 5b). The predictor and events are identical in the two analyses.

No participant’s 95% prediction interval for time to onset reached an upper limit within follow-up (Figure 5c): the model cannot say when, or whether, any individual will develop symptoms.

### Common-Trajectory Assumption

The clock assumes that individuals with the same %p-tau217 level follow the same trajectory. A single observed %p-tau217 value, the quantity available from one blood test, explained 6% of the between-person variance in the rate of change in the ADNI clock-development sample (adjusted R² 0.06, 95% CI 0.02 to 0.10; Figure 6a). Substituting the model-based value at the midpoint of follow-up, which incorporates the participant’s own rate of change, raised the explained variance to 17% (adjusted R² 0.17, 95% CI 0.11 to 0.24; Figure 6b).

**Figure 6.**
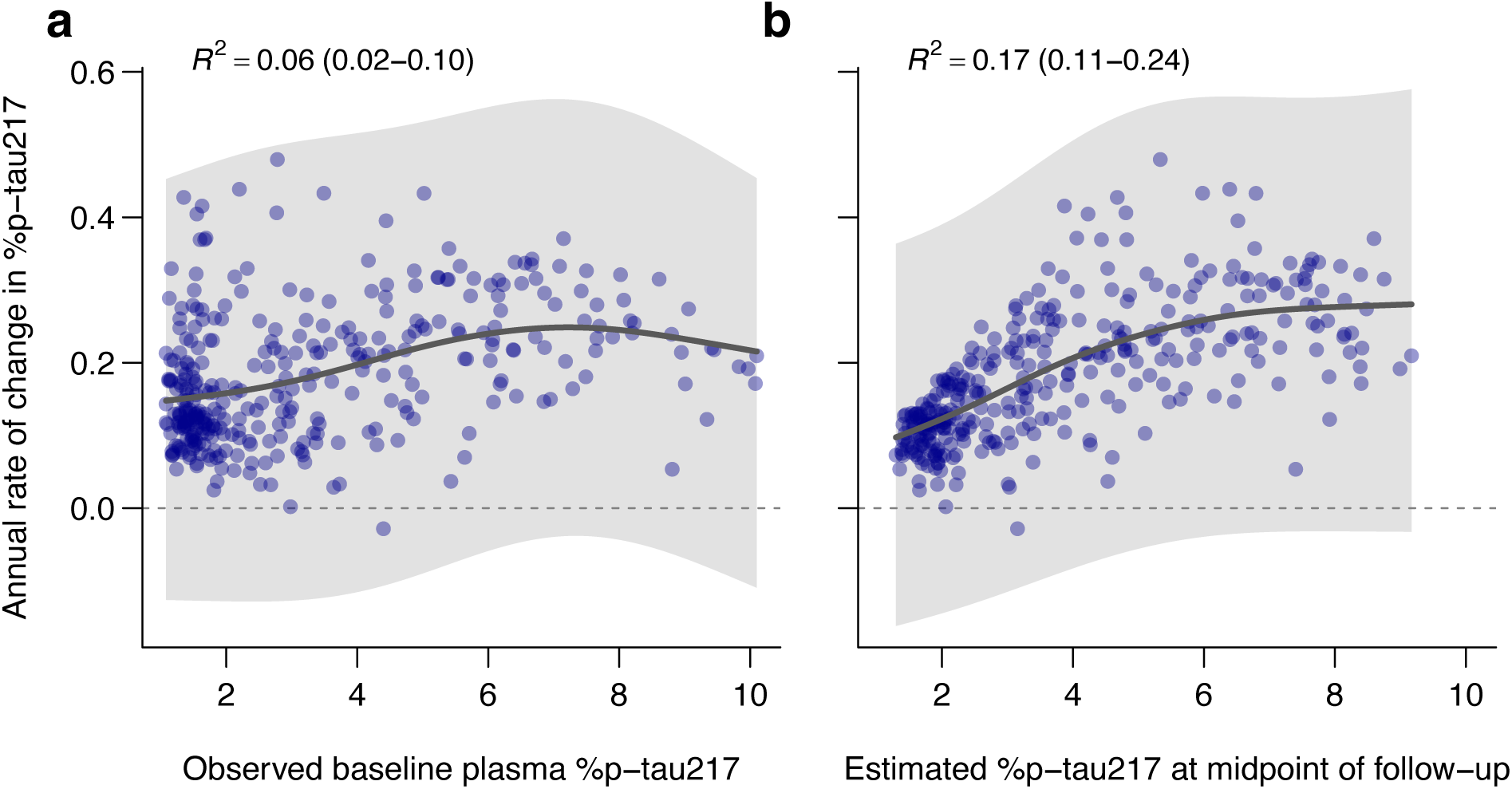
Annual rate of change in plasma %p-tau217 as a function of %p-tau217 level in the ADNI clock-development sample (n=345). Points are participant-specific rates from a linear mixed model with random intercepts and slopes. The curve is the mean rate as a nonlinear function of level, and the shaded band is the 95% prediction interval for an individual’s rate. R² is the mean adjusted R² across resamples, with its 2.5th–97.5th percentile interval. In (a), level is the observed value at the first plasma collection, the quantity available from a single blood test. In (b), level is the model-based value at the midpoint of follow-up, which incorporates the participant’s own rate of change. The dashed line marks a rate of zero.

Consider two participants, both with a %p-tau217 of 3.0, just below the positivity threshold of 4.06. The 95% prediction interval for their rates at that level runs from below zero to roughly 0.5 per year: one may cross the threshold within a couple of years, while the other may never cross it at all, yet the clock assigns them the same estimated time to positivity.

Under either definition of level, the overwhelming majority of the variance in rate is left unexplained.

## Discussion

A reliable way to forecast when symptoms will begin is among the central goals of Alzheimer research. A single blood test that could do so for a cognitively unimpaired person would change how we counsel patients, time treatment, and design trials. The reported accuracy drew attention accordingly. But the accuracy is a property of the design, not of the biomarker: the models do not predict age at symptom onset from a biomarker; they predict age from age.

Every primary analysis contains both design artifacts: the same quantity enters the predictor and the outcome, and the outcome is confined to a region fixed by the predictor. Together they account for nearly all of the reported result. Either alone is sufficient to produce the association, and replacing the biomarker with a random number reproduced it in nearly every simulation, accounting for most of its magnitude in Knight ADRC and exceeding it in ADNI. Once both are accounted for, the biomarker explains essentially none of the remaining variation, and the raw measurement outperforms the clock.

A follow-up preprint by three of the original authors reports that the clock exceeds a randomization null.^19^ That null shuffled estimated age at positivity across participants. This discards the design constraints: the estimate must fall within a window around age at blood draw and must precede the participant’s own onset. As a result, the null understates what the design alone produces, with a mean adjusted R² near zero.

The second reported conclusion is that time from positivity to onset is shorter in older individuals. This also follows from the shared quantity and the bound, and the random comparator reproduces it. Zhu et al^15^ independently showed that this analysis, which they identify in Petersen et al, produces a negative association by construction. Under the null, it gave false-positive rates as high as 100%, so it cannot serve as a test of association. They also identified collider bias from restricting the sample to participants whose estimated positivity preceded onset. The corrected analysis by Zhu et al estimates hazard at a given age, not the interval from positivity to onset. When we removed the shared quantity and the bound, estimated age at positivity was not associated with time to onset. No analysis, theirs or ours, supports a shorter onset interval at older ages.

The bound does not disappear between the analysis of age at onset and the analysis of time from positivity to onset; it relocates. For age at onset, the outcome is constrained relative to age at blood draw by the follow-up window. For time from positivity to onset, the outcome is constrained relative to estimated age at positivity, because it is constructed as age at onset minus that quantity. The constraint in the second analysis is therefore not the constraint in the first, and the recent defense that duration is unassociated with baseline age^6^ is irrelevant.

Independence from age at blood draw is expected under the artifact; an association with positivity age alongside no association with age at blood draw is precisely the pattern the structural account predicts. The same construction is common in the disease clock literature. A typical example regresses the time from amyloid to tau positivity on age at amyloid positivity. Because tau positivity must follow amyloid positivity within the observed age range, the longest possible interval shrinks as age at amyloid positivity increases. The design alone therefore produces a negative association, even when the interval does not depend on age.

The survival analysis shows the same dependence directly. The age-scale model in the original report enters each participant into the risk set at birth, although no event could be counted before the blood draw or before estimated positivity. Participants with later positivity ages are therefore credited with more symptom-free time, which is an immortal time bias.^17,18^ The analysis of Zhu et al avoids this through left truncation. With the immortal time removed, the same events show no association, and the model cannot give any individual an upper limit on when symptoms might begin, or rule out that they never will.

Even if the clock carried prognostic biomarker information, the model could not be deployed clinically as presented. Built only in people already known to have progressed, it neither attempts nor is able to identify who will progress, so every predicted age at onset carries an implicit qualification: “assuming this individual will progress.” A model that cannot answer the *if* cannot meaningfully answer the *when*.

Two further problems, not examined in our analyses, bear directly on the headline claim. First, although the claim is that a single blood test can time symptom onset, every analysis draws on repeated measurements wherever they exist: participants with more than one plasma sample were assigned the average of the estimates from all their samples,^9^ so the reported accuracy reflects longitudinal data, not a single draw, and how well one measurement alone predicts onset is never shown. Second, the headline R² values come from regressions refit within each cohort and evaluated on the same participants used to fit them. They show how well a model can be made to fit outcomes already observed, not how accurately it would predict onset for a new patient, and so do not validate a prediction model.

At a more fundamental level, the framework also rests on an assumption the data do not support, that everyone follows a common trajectory. In the development data, a single measurement explains only 6% of the between-person variance in the rate of accumulation. The developers’ own simulations identify this between-person variation in rate as the dominant source of error in the clock,^20^ so mapping a level to a time through a group-mean curve adds error without adding information.

An adjusted R² of 0.6, for a clinical outcome from a single blood draw, is a magnitude that should invite scrutiny. Age at symptom onset is derived from the CDR, which is least reliable at the transition that defines onset, where agreement on CDR 0.5 is only 66%^21^ and chance-corrected reliability is poor (κ = 0.33).^22^ Test-retest reliability of plasma %p-tau217 is approximately 0.90.^23^ Measurement error in both variables places a ceiling on observable R² well below 1, and beneath that ceiling the variance is shared with cognitive reserve, cerebrovascular disease, Lewy body pathology, TDP-43, and neurodegeneration.^24,25^ Implausibly large effects have long signaled design issues rather than discovery.^13^

This study has limitations. The progressor analyses inherit the small samples of the provided data, particularly in ADNI. The common-trajectory analysis was conducted in ADNI only, and the survival analysis, one cohort with 30 events, establishes the timescale dependence but not the magnitude of any true association.

The central claim that a single blood test can predict the age of symptom onset with median accuracy of less than 4 years does not hold. Findings of this kind redirect research effort toward refining a method that only adds noise to the raw signal, promise trial enrichment that the model cannot deliver, and, through a live web application and direct-to-consumer p-tau217 testing, reach individuals who will be told when they can expect symptoms of Alzheimer’s disease.

Predicting clinical change with the accuracy claimed for these clocks is exactly where the field would like to be, and blood-based markers may yet take us there. The evidence does not show that we have arrived. Clinical prognosis requires estimating both the probability of progression and the uncertainty surrounding its timing. A disease clock that assumes a common trajectory for everyone provides neither.

## Data Availability

ADNI data are available to qualified researchers at adni.loni.usc.edu. Knight ADRC progressor data were shared by the authors of the original report and are available from them on request.

https://github.com/mcdonohue/blog/tree/main/posts/design-artifacts

## Acknowledgements

We thank the authors of the original report for sharing the data used in the progressor analyses and for their engagement in discussions of these issues.

Data from the Knight Alzheimer Disease Research Center were provided by the authors of the original report and were collected with support from NIA grants P30AG066444, P01AG003991 and P01AG026276.

Data collection and sharing for the Alzheimer’s Disease Neuroimaging Initiative (ADNI) is funded by the National Institute on Aging (National Institutes of Health Grant U19AG024904). The grantee organization is the Northern California Institute for Research and Education. In the past, ADNI has also received funding from the National Institute of Biomedical Imaging and Bioengineering, the Canadian Institutes of Health Research, and private sector contributions through the Foundation for the National Institutes of Health (FNIH) including generous contributions from the following: AbbVie, Alzheimer’s Association; Alzheimer’s Drug Discovery Foundation; Araclon Biotech; BioClinica, Inc.; Biogen; Bristol-Myers Squibb Company; CereSpir, Inc.; Cogstate; Eisai Inc.; Elan Pharmaceuticals, Inc.; Eli Lilly and Company; EuroImmun; F. Hoffmann-La Roche Ltd and its affiliated company Genentech, Inc.; Fujirebio; GE Healthcare; IXICO Ltd.; Janssen Alzheimer Immunotherapy Research & Development, LLC.; Johnson & Johnson Pharmaceutical Research & Development LLC.; Lumosity; Lundbeck; Merck & Co., Inc.; Meso Scale Diagnostics, LLC.; NeuroRx Research; Neurotrack Technologies; Novartis Pharmaceuticals Corporation; Pfizer Inc.; Piramal Imaging; Servier; Takeda Pharmaceutical Company; and Transition Therapeutics.

Data used in preparation of this article were obtained from the Alzheimer’s Disease Neuroimaging Initiative (ADNI) database (adni.loni.usc.edu). As such, the investigators within the ADNI contributed to the design and implementation of ADNI and/or provided data but did not participate in analysis or writing of this report. A complete listing of ADNI investigators can be found at: http://adni.loni.usc.edu/wp-content/uploads/how_to_apply/ADNI_Acknowledgement_List.pdf.

## AI Use

The authors used Claude Opus 5 (Anthropic) for copyediting, wording, and comments on code. All content was reviewed and revised by the authors, who take full responsibility for it.

## Ethical approval

This study used deidentified data from ADNI and the Knight ADRC, whose protocols were approved by the institutional review boards of all participating sites, with written informed consent from all participants. This secondary analysis of deidentified data did not require additional ethical approval.

## Data sharing

ADNI data are available to qualified researchers at adni.loni.usc.edu. Knight ADRC progressor data were shared by the authors of the original report and are available from them on request. Analysis code is available at https://github.com/mcdonohue/blog/tree/main/posts/design-artifacts

**Supplementary Material**

**Supplementary Methods**

**Supplementary Figures**

## Supplementary Methods

### Statistical Analysis

In each analysis, the aim was to separate the association attributable to biomarker signal from the association produced by two design features: a shared component, in which the same quantity enters both the predictor and the outcome, and a bound, in which the outcome is confined to a region fixed by the predictor.

We decomposed the clock into age at blood draw, defined as age at the first plasma collection, and estimated time from %p-tau217 positivity (time from positivity). In the first set of analyses, age at symptom onset was regressed, using ordinary least squares (OLS), on age at blood draw, on time from positivity, and on the clock-estimated age at positivity, each separately. As a null comparison, we replaced time from positivity with a random value drawn uniformly over the same range and repeated this 10,000 times to generate a distribution of adjusted R² expected from study design constraints alone, in the absence of any biomarker signal. We further compared the clock against a model using age at blood draw and raw %p-tau217 level directly. Models were compared by adjusted R² and by the Akaike information criterion (AIC).^1^

In a sensitivity analysis, we excluded participants whose symptom onset preceded the blood draw and repeated the comparisons of the clock, age at blood draw, and age at blood draw with raw %p-tau217. The clock is proposed to predict when symptoms will begin in people who are cognitively unimpaired. For a participant whose blood was drawn after onset, the measurement comes from someone already impaired, and the onset it is used to predict has already occurred.

In a second set of analyses, we removed age at blood draw from the outcome by regressing time from blood draw to symptom onset on estimated time from positivity, obtained from TIRA, from SILA, and from a random value drawn uniformly over the same range. Age at blood draw appears on neither side of this regression, so the shared component is absent, but the bound remains.

Because onset was required to follow estimated positivity, a participant not yet positive at the draw could be included only if onset occurred after the time at which the clock places positivity. For these participants, estimated time from positivity sets a floor on time to onset, and the more negative the estimated time from positivity, the higher the floor. For participants already positive at the draw (estimated time from positivity greater than 0), the floor falls before the draw and places little constraint on the observed time to onset. We therefore repeated the analysis restricted to participants already positive at the draw. If the association reflects biomarker information, it should persist in this group; if it reflects the bound, it should largely disappear.

In a third set of analyses, time from estimated %p-tau217 positivity to symptom onset (observed age at onset minus estimated age at positivity) was regressed on estimated age at %p-tau217 positivity, based on TIRA, SILA, and a random value. Because the outcome is age at onset minus the predictor, the covariance between them equals the covariance between age at onset and the predictor minus the variance of the predictor, and is negative even when the biomarker carries no information.^2^ The outcome is also bounded by the predictor, because observed ages at onset fall within a limited range.

In a fourth set of analyses, we included nonprogressors and used survival analysis to test whether the association depends on the shared component and the bound. Interval-censored proportional hazards models, fit with the software used in the original report (icenReg,^3^ semiparametric baseline hazard), regressed symptom onset on estimated age at positivity on two timescales. The first was age, as in the original report, in which age at blood draw enters both the predictor and the outcome and every participant contributes follow-up from birth. The second was time from the blood draw, or from the estimated time of positivity for participants not yet positive at the draw. Because onset was counted as an event only after estimated positivity, earlier follow-up is immortal time,^4,5^ during which the event, as defined, cannot occur. Confidence intervals were obtained by bootstrap resampling (5,000 samples), and individual 95% prediction intervals for time to symptom onset were derived from the fitted survival curves. Survival curves are shown until fewer than five participants remained under observation.

In a fifth set of analyses, we evaluated the common-trajectory assumption underlying the clock models. Petersen et al state that once amyloid plaques and neurofibrillary tangles begin to accumulate, the burden of pathology “follows a remarkably consistent trajectory across individuals.”^6^ Under this assumption, individuals at the same level of pathology progress at the same rate, so a person’s rate of %p-tau217 accumulation is determined by their current level, and a single value should explain most of the between-person variance in the rate of change.

To quantify how much of that variance one measurement explains, we fit a linear mixed model with random intercepts and slopes to longitudinal %p-tau217 in the ADNI clock-development sample (n=345), with years since the first plasma collection as the time scale. The variances of the random intercept and slope and their correlation were estimated freely, assuming independent residuals and constant variance given the random effects. We resampled each participant’s intercept and slope 2,000 times from their conditional distribution given the observed data and the estimated variance components. For each resample, we regressed the rate on the participant’s %p-tau217 level using a penalized regression spline. We used two definitions of level: the observed value at the first plasma collection, the quantity available from a single blood test, and the model-based value at the midpoint of follow-up, which incorporates the participant’s rate of change. We report the mean adjusted R² and the mean fitted curve across resamples, with the 2.5th and 97.5th percentiles of the adjusted R² as a 95% confidence interval. The 95% prediction interval for an individual’s rate at a given level was obtained by fitting conditional quantiles to the pooled resamples using quantile additive regression. Age at %p-tau217 positivity was estimated with the SILA algorithm in the R package silaR. Analyses used lme4, mgcv, qgam and icenReg in R, version 4.6.0.

## Supplementary Figures

**Supplementary Figure S1.**
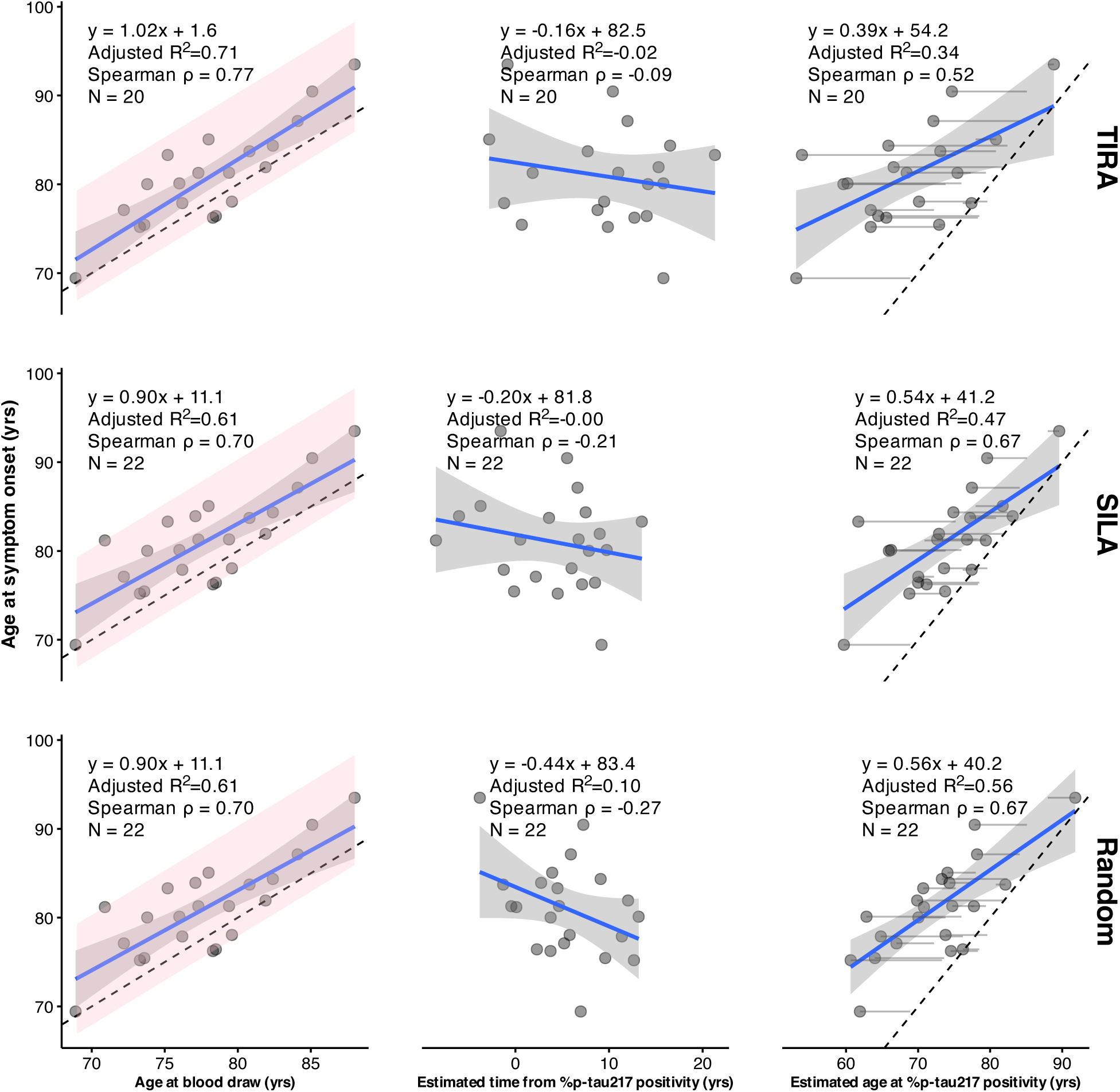
Relationships in ADNI between age at symptom onset and three predictors: age at blood draw (first plasma collection), estimated time from %p-tau217 positivity, and their difference, age at blood draw minus estimated time from %p-tau217 positivity (ie, estimated age at %p-tau217 positivity). Time from %p-tau217 positivity is estimated by the TIRA method (top row), the SILA method (middle row), or by randomly sampling from a uniform distribution over the observed range of estimates (bottom row). Panels include fitted linear regression lines with 95% confidence bands, the regression equation, adjusted R², Spearman ρ, N, and a dashed identity line. In the rightmost panels, faint grey lines connect each person’s age at blood draw to their estimated age at %p-tau217 positivity. The pink regions span the observed window of onset, from 2.1 years before to 10.3 years after the blood draw.

**Supplementary Figure S2.**
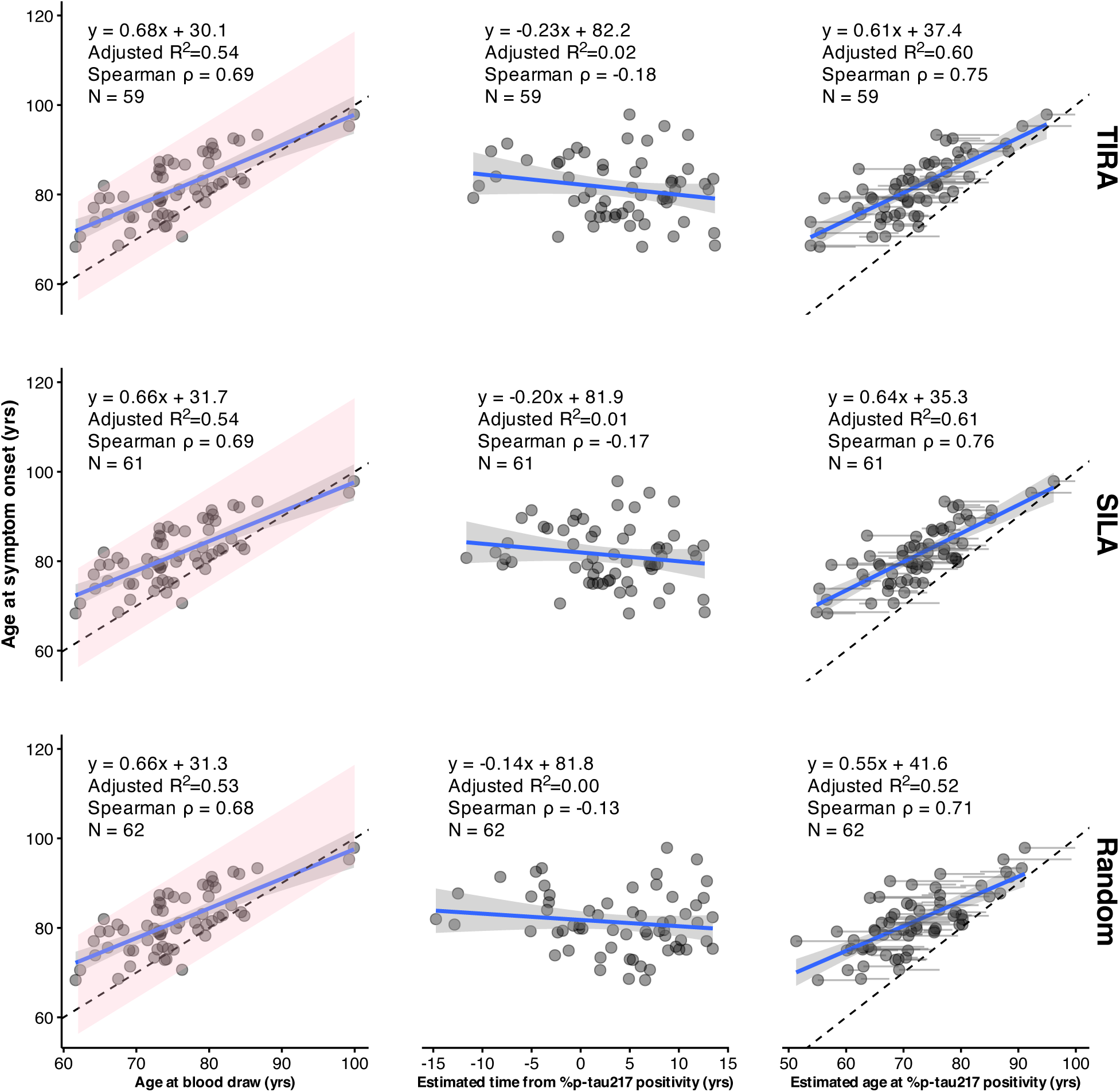
Relationships in Knight ADRC between age at symptom onset and three predictors: age at blood draw (first plasma collection), estimated time from %p-tau217 positivity, and their difference, age at blood draw minus estimated time from %p-tau217 positivity (ie, estimated age at %p-tau217 positivity). Time from %p-tau217 positivity is estimated by the TIRA method (top row), the SILA method (middle row), or by randomly sampling from a uniform distribution over the observed range of estimates (bottom row). Panels include fitted linear regression lines with 95% confidence bands, the regression equation, adjusted R², Spearman ρ, N, and a dashed identity line. In the rightmost panels, faint grey lines connect each person’s age at blood draw to their estimated age at %p-tau217 positivity. The pink regions span the observed window of onset, from 5.7 years before to 16.4 years after the blood draw.

**Supplementary Figure S3.**
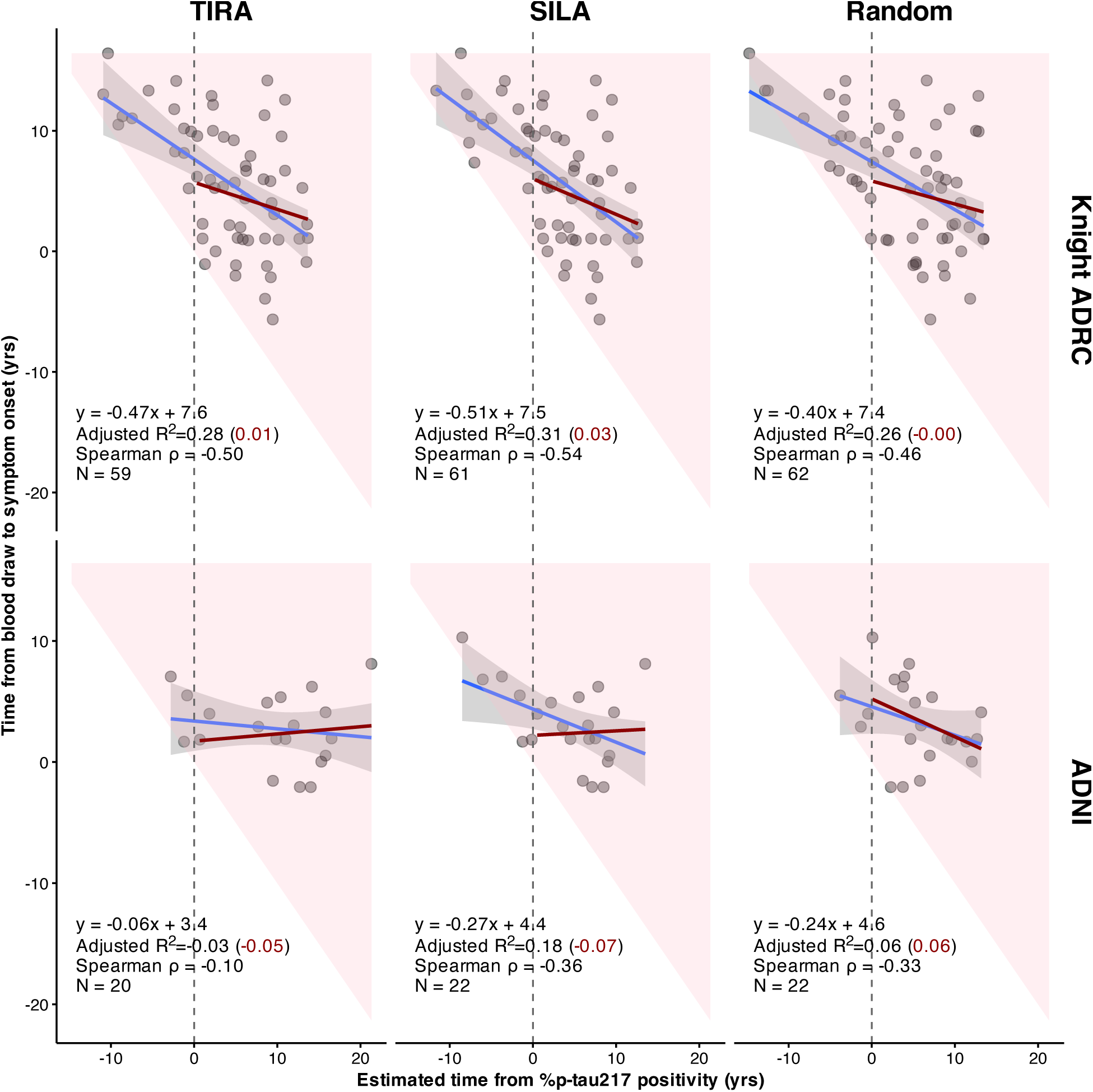
Time from blood draw to symptom onset versus estimated time from %p-tau217 positivity in Knight ADRC (top row) and ADNI (bottom row), with time from positivity estimated by the TIRA method (left), the SILA method (middle), or drawn uniformly over the observed range of estimates (right). Negative values of estimated time from positivity indicate participants not yet positive at the blood draw (left of the dashed vertical line). The pink region shows the values permitted by the design: because onset must follow estimated positivity, time to onset cannot fall below the diagonal. Blue lines show linear fits in all participants, with 95% confidence bands; dark red lines show fits restricted to participants already positive at the blood draw. Each panel reports the regression equation, adjusted R² for all participants (restricted sample in parentheses), Spearman ρ, and N.

**Supplementary Figure S4.**
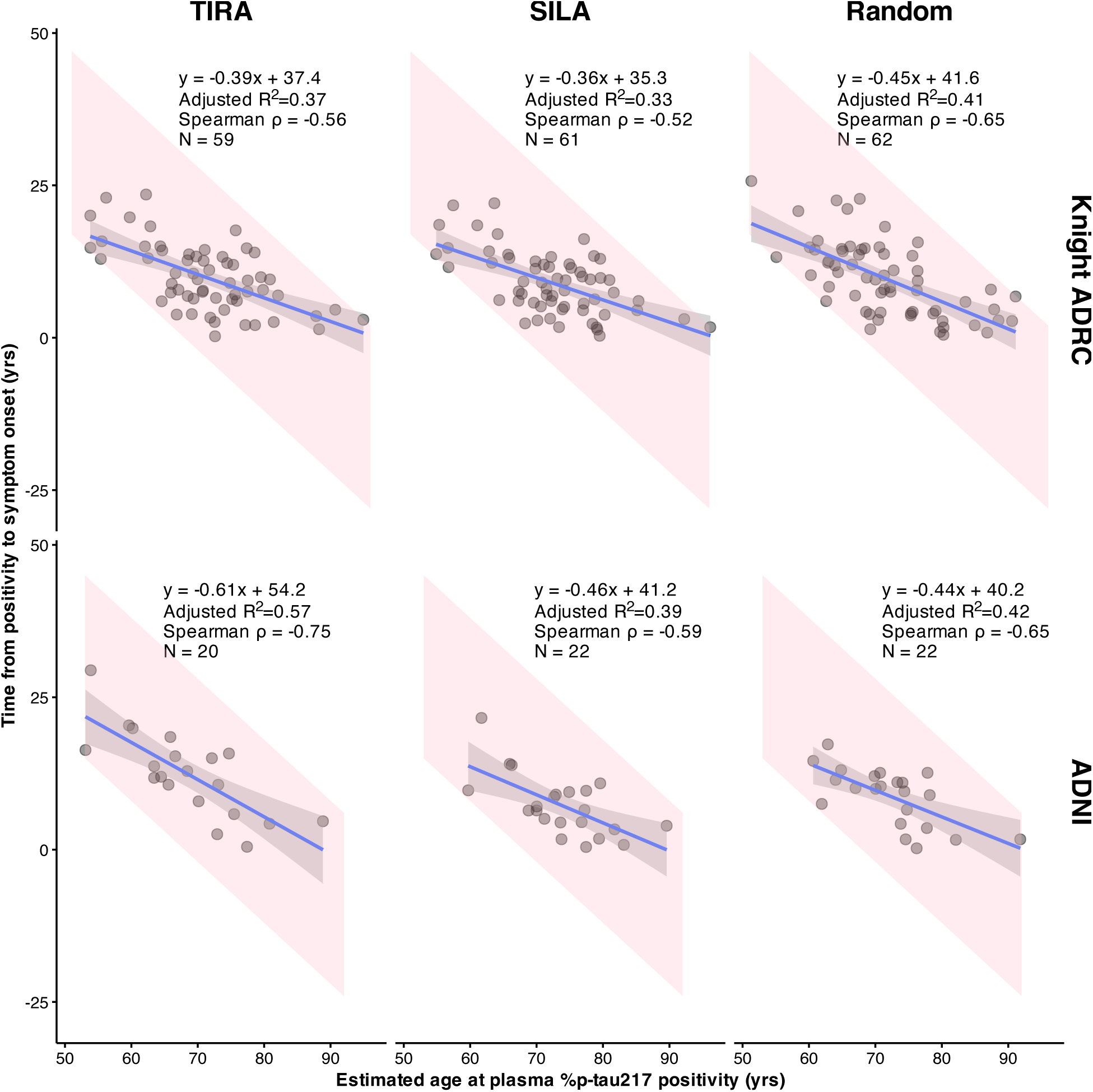
Time from estimated %p-tau217 positivity to symptom onset (age at symptom onset minus estimated age at positivity) versus estimated age at positivity in Knight ADRC (top row) and ADNI (bottom row), with age at positivity estimated by the TIRA method (left), the SILA method (middle), or as age at blood draw minus a random time drawn uniformly over the observed range of estimates (right). Panels include fitted linear regression lines with 95% confidence bands, the regression equation, adjusted R², Spearman ρ, and N. The pink regions show the values possible given the observed range of ages at symptom onset in each cohort: the older the estimated age at positivity, the shorter the duration can be.

